# Protocol Study for the Development of *MINDS* (Mental Health and Intervention for New Doctors and Suicide Prevention) in Mexico

**DOI:** 10.64898/2025.12.04.25341658

**Authors:** Pamela Espinosa Méndez, Abril Téllez Buendía, Rocío Jurado Galicia, Carlos Alfonso Tovilla-Zárate, Thelma Beatriz Gonzales-Castro, Humberto Nicolini, Martha Catalina Pérez Gonzales, Francisco Gutiérrez Rodríguez, Janitza L. Montalvo-Ortiz, José Jaime Martínez-Magaña, Alma Delia Genis-Mendoza

**Affiliations:** Comisión Nacional de Salud Mental y Adicciones (CONASAMA), Secretaría de Salud, CDMX, México; División Académica Multidisciplinaria de Comalcalco, Universidad Juárez Autónoma de Tabasco, Comalcalco, Tabasco; División Académica Multidisciplinaria de Jalpa de Méndez, Universidad Juárez Autónoma de Tabasco, Jalpa de Méndez, Tabasco; Laboratorio de Genómica de Enfermedades Psiquiátricas y Neurodegenerativas, Instituto Nacional de Medicina Genómica, CDMX, México; Department of Psychiatry, Yale School of Medicine, New Haven, CT, USA; Veterans Affairs Connecticut Healthcare System, West Haven, CT, USA; Hospital Psiquiátrico Infantil Juan N. Navarro, Secretaría de Salud, CDMX, México

**Author notes:** Corresponding to: José Jaime Martínez-Magaña ( /); VA Connecticut Healthcare Errera Community Care Center - Orange Annex, 200 Edison Road, Orange, CT 06477. Alma Delia Genis-Mendoza ( /); Instituto Nacional de Medicina Genómica, Periferico Sur 4809, Arenal Tepepan, Tlalpan, 14610 Ciudad de México, CDMX, Mexico.

**Keywords:** Mental Health, Suicide, Medical students, Biomarkers, Primary Prevention, Longitudinal Studies

## Abstract

Addressing mental health problems is a global public health priority. Healthcare trainees are the future professionals responsible for the health of the population, and they also face significant mental health challenges that need our support. Over the past decade, the prevalence of mental health problems among healthcare trainees has increased, highlighting the urgent need to care for those who will care for others. In response, as part of the National Program for Suicide Prevention in Mexico and to address the mental health problems in healthcare trainees in Mexico, we developed ***MINDS*** (**M**ental Health and **In**tervention for **N**ew **D**octors and **S**uicide Prevention), a program designed to evaluate, prevent, and treat mental health problems in healthcare trainees, with a particular focus on suicide risk. MINDS targets students in undergraduate programs who are participating in medical internships and social service programs across all states of Mexico. Here, we aimed to present the MINDS program protocol. The program follows a longitudinal design, beginning with an initial assessment to identify trainees at risk and referring them to clinical treatment as needed. At this stage, participants will also provide a blood or saliva sample to create a DNA biorepository. A follow-up evaluation will occur six months later. MINDS assesses a wide range of mental health domains, including depression, anxiety, substance use disorders, eating behaviors, and ***suicide risk***. By generating critical data and a practical framework for the mental health of medical trainees, MINDS will promote well-being and contribute to a healthier environment within the medical profession in Mexico.

## Introduction

According to the Global Burden of Disease, mental health problems, led by depression and anxiety, pose a significant global public health concern [1, 2], which has been further exacerbated by the COVID-19 pandemic [3]. Among healthcare professionals, these issues may be even more pronounced, with substantial increases in mental health problems, including elevated *suicide risk* [4]. Hypotheses suggest that this rise may be driven by burnout [5], highlighting the urgent need for strategies that promote better mental health among healthcare professionals. Early interventions have primarily targeted emotional health, focusing on psychological support, stress management, resilience training, and development of communication skills [4]. However, other approaches recognize the importance of combining individualized strategies with systemic and institutional changes, addressing organizational factors and avoiding the stigmatization of healthcare professionals as weak or incompetent [6, 7]. Despite these efforts, effectively improving mental health and reducing burnout among healthcare professionals remains a significant challenge.

Studies suggest that mental health problems among healthcare professionals, particularly *physicians*, often begin in the early stages of their academic training. Evidence indicates that medical students are at higher risk of developing mental health problems compared with students in other fields [8]. Meta-analyses had estimated that the prevalence of depression among medical students and resident physicians is higher than in the general population, with rates of approximately 27.2% and 28.8%, respectively[9, 10]. Notably, an analysis of seven longitudinal studies found that 15.85% of the increase in depressive symptoms over time was directly associated with the transition into residency programs [9]. The growing responsibility of caring for large numbers of patients across multiple levels of care, combined with the academic demands imposed by universities, places a significant burden on healthcare trainees, potentially leading to burnout [11–13]. This dual challenge of clinical workload and academic rigor, coupled with the emotional demands of direct patient care, creates particularly stressful conditions for medical trainees. The increased prevalence of mental health problems in medical trainees underscores the global importance of coordinated systemic interventions to safeguard the well-being of future physicians.

In alignment with the National Program for Suicide Prevention in Mexico, which prioritizes mental health and well-being, including medical trainees, we developed **MINDS** (*Mental Health and Intervention for New Doctors and Suicide Prevention*). MINDS is designed to implement a comprehensive approach for evaluating, preventing, and treating mental health problems and *suicide risk* in medical trainees. The program incorporates digitalized, internationally validated instruments accessible via smartphone, computer, or tablet, ensuring both scientific rigor and practical feasibility while potentially reducing the risk of severe mental health outcomes. In addition, we will establish a DNA repository to enable integration of biological data and the identification of potential novel biomarkers for mental health problems, not only in medical trainees but in the Mexican population. In this paper, we present the MINDS study protocol. The strategies outlined in the MINDS protocol are expected to provide critical guidance for future prevention strategies and intervention efforts, ultimately advancing the well-being of medical trainees across Mexico.

## Materials and Methods

### Participants

This manuscript presents MINDS, a Mexican national initiative structured around three core components: ***evaluation, prevention, and treatment***. We aim to address mental health problems and suicide risk among medical trainees in Mexico (**Figure 1**). Through collaboration with the country’s principal academic and healthcare training programs, and in coordination with educational and health authorities, undergraduate medical interns, and social service trainees will be invited to participate. We estimate participation from over 76 universities across all 32 states of Mexico ***[14]*.**

**Figure 1.**
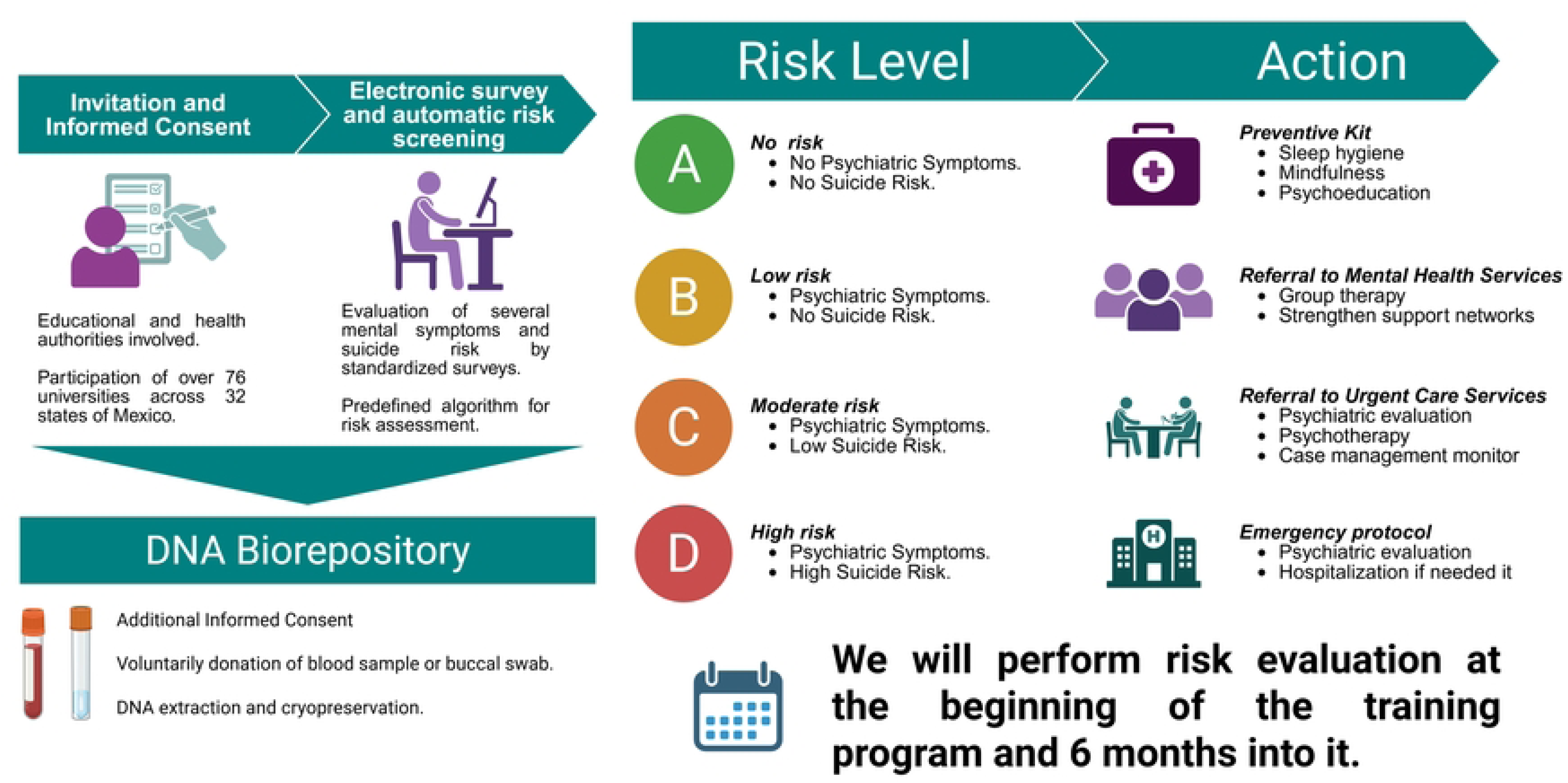
**Overview of MINDS (M**ental Health and **In**tervention for **N**ew **D>octors and Suicide Prevention)**

### Procedures

#### Evaluation

We will conduct a first evaluation at the beginning of the trainees’ program using a digital platform accessible via computer or smartphone. MINDS will start on *March 2026,* and we plan to keep the program ongoing until *March of 2030*. This digital assessment will include validated instruments designed to evaluate anxiety, depression, substance use, eating behaviors, psychiatric symptoms, and suicide risk. Participants will access the Screening Questionnaire Platform, where they will first review an electronic informed consent form. The consent forms will clearly state that participation in MINDS is entirely voluntary. That refusal to participate or withdrawal from the study will not affect the trainee’s academic standing, evaluation, or progression in their medical program. We will not include trainees who are not formally enrolled in their educational programs at the time of the first evaluation, or those without access to the digital evaluation forms. The protocol has been approved by the Research Committee of the Fray Bernardino Álvarez Psychiatric Hospital (HPFBA, number: CI-1138).

The initial evaluation will include the following instruments:

##### Sociodemographic Data Sheet

Includes contact information, age, gender, academic year, state where the internship or service is being conducted, name of the unit where the participant is completing the undergraduate internship or social service, university where the medical degree is being pursued, name of the Head of Teaching at the service unit, marital status, type of housing, number of financially dependent individuals, travel time from home to the service unit, and the hospital service currently assigned for rotation.

##### Mental Disorders Screener Questionnaire [15]

An instrument designed to identify symptoms of 13 mental disorders in the population.

##### Depression

The *Patient Health Questionnaire 9-item scale (PHQ-9)* [16]. The PHQ-9 is a nine-item questionnaire used to screen for and assess the severity of depression. It is widely used in both clinical and research settings and directly reflects the diagnostic criteria for Major Depressive Disorder (MDD).

##### Anxiety

The *Generalized Anxiety Disorder 7-item scale (GAD-7)* [17]. The GAD-7 is a brief, self-report questionnaire used to assess the severity of anxiety symptoms. It consists of seven questions that gauge the frequency of various anxiety symptoms over the past two weeks. Each item is scored on a 4-point scale, and the total score (ranging from 0 to 21) indicates the severity of anxiety.

##### Substance Use

The *World Health Organization (WHO) Alcohol, Smoking and Substance Involvement Screening Test (ASSIST)* [18]. The ASSIST is a questionnaire that screens for different levels of problematic or risky substance use. It includes three subscales: alcohol, tobacco, and other psychoactive substances. Each subscale provides information on lifetime and past-trimester use of each substance, categorizing individuals into three risk levels: low (no intervention needed), moderate (behavioral intervention required), and high (immediate treatment required).

##### Suicide Risk

The short version of the *Columbia–Suicidal Ideation Severity Rating Scale (C-SSRS)* [19]. The C-SSRS is a widely validated and commonly used tool for assessing the risk of suicidal behavior. It serves as an initial screening tool to categorize individuals into low-, moderate-, or high-risk groups. In the screener version, answering “yes” to questions 1 or 2 indicates low risk; answering “yes” to question 3 suggests moderate risk; and answering “yes” to questions 4, 5, or 6 indicates high risk.

##### Eating Disorders

The *Eating Disorder Examination Questionnaire (EDE-Q)* [20]. The EDE-Q assesses behaviors associated with eating disorders and includes four subscales: Dieting and Restrictive Behaviors, Eating Preoccupation, Weight Preoccupation, and Shape Preoccupation. A score higher than 1.09 has been shown to identify individuals at risk of eating disorders.

#### Automatic evaluation system

We will develop a risk evaluation system within the digital platform to identify medical trainees at high risk for mental health problems and refer them to specialized local healthcare services for appropriate treatment. Using validated algorithms from the PHQ-9, GAD-7, ASSIST, EDE-Q, and C-SSRS surveys, the system will stratify participants into four risk groups based on the criteria outlined in **Table 1**. Each trainee will receive an encrypted, personalized email summarizing their results and providing tailored information and links to available support and mental health care resources. A follow-up evaluation will be conducted *six months* after the initial assessment, including both those who participated in the baseline evaluation and those who initially declined. This longitudinal design will allow us to monitor symptom trajectories, evaluate the effectiveness of our interventions, and identify new-onset cases emerging during the course of training.

**Table 1.**
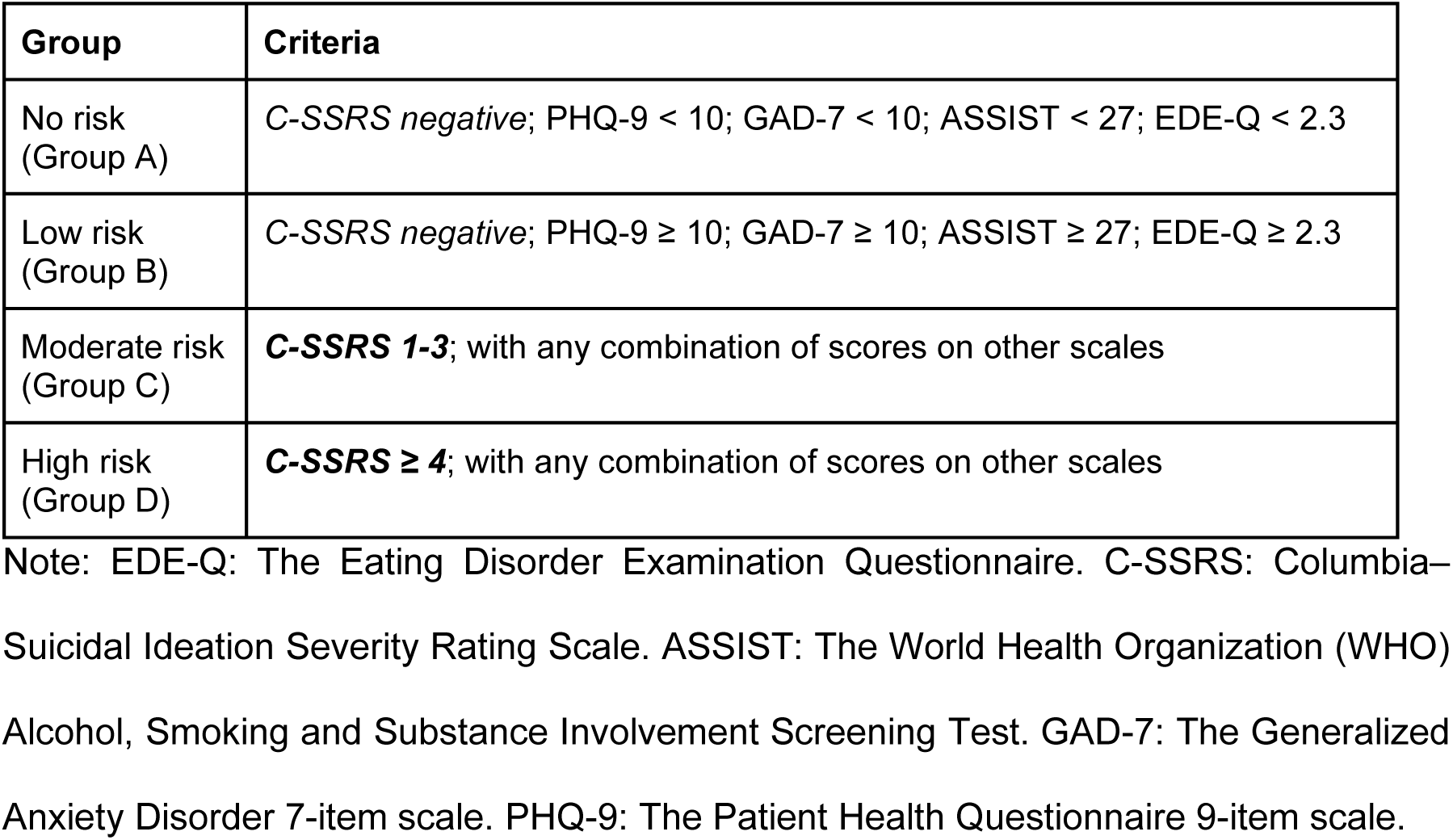
Criteria for the assignment of each trainee into groups.

#### Referral of Trainees At-Risk

Following the initial evaluation, we will determine each trainee’s risk of psychiatric disorders and promptly notify them of their results. The digital platform will communicate the risk level to the trainees, the corresponding university teaching department, and the state authority responsible for medical trainees (**Figure 2**), ensuring coordinated care for those at risk. This integrated approach will enable timely referral to mental health services, with all referrals monitored by a designated case manager appointed by each state’s Mental Health and Addictions Coordinator. Importantly, all consent forms and procedures will emphasize that participation in the MINDS program and the results of the risk assessment will not influence any future academic decisions regarding the trainee.

**Figure 2.**
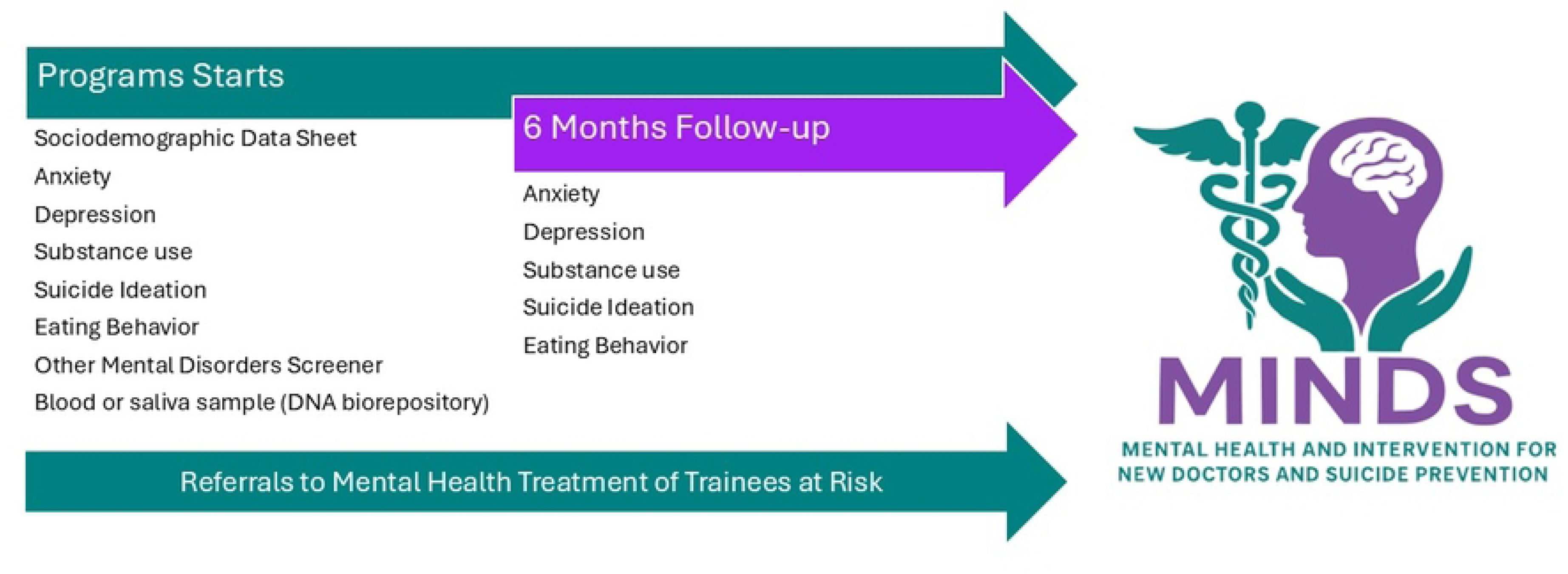
**Overview of MINDS (M**ental Health and Intervention for **N**ew **D**octors and Suicide Prevention) **steps**.

Trainees identified as not at risk (Group A) will also receive their results, accompanied by an informative brochure containing preventive recommendations for maintaining mental well-being, including guidance on sleep hygiene, mindfulness practices, and psychoeducation. These trainees will be invited to participate in preventive group activities organized by their institutions and will be provided with information on accessing mental health services should future needs arise.

For trainees identified as low to high risk (Group B to D), referrals will be made to the corresponding state mental health services within three to five days, following a severity-based protocol:

Low risk (Group B): The teaching department will notify the case manager, who will arrange referrals to first-level care services for early intervention. These may include group sessions focused on stress management and strengthening social support networks.

Moderate risk (Group C): The case manager will coordinate with the teaching department for urgent referral (≤72 hours) to second-level care (general hospitals with psychiatry services). The trainee will receive brief psychotherapy and psychiatric evaluation, with ongoing monitoring by the case manager.

High risk (Group D): The teaching department will activate the emergency protocol immediately. The case manager will notify the trainee’s family and relevant health authorities to ensure immediate transfer (*<24 hours*) to emergency or psychiatric hospital services for evaluation and possible hospitalization.

### DNA Biorepository

Most mental disorders exhibit significant heritability [21]. Genomic studies, including genome-wide association studies (GWAS), have identified thousands of associated genomic loci [22–24]. Using GWAS data, novel tools known as polygenic risk scores (PRS) or polygenic scores, have emerged as potential biomarkers for mental disorders [25]. A PRS is a single numerical value that estimates an individual’s overall genetic risk for a specific trait, such as mental health disorder. It is calculated by summing the effects of genetic variants across the genome, weighted by their estimated effect sizes from GWAS. PRS has shown promise as a tool for risk assessment, potentially helping to identify individuals who are more likely to develop mental health conditions [26].

As a separate aim, we will invite trainees to participate in a *genomic sub-study*, in which a blood or saliva sample will be collected at the time of the first evaluation for DNA extraction and long-term storage for potential genotyping in future studies. The samples will be stored in the biorepository of the Instituto de Medicina Genómica (INMEGEN) for future analysis. A second informed consent will be provided for this genomic component. All informed consent forms will clearly state that participation in MINDS is entirely voluntary. That refusal to participate or withdrawal from the study will not affect the trainee’s academic standing, evaluation, or progression in their medical program. Additionally, the informed consent will specify that the collected biological samples may be used for future research.

### Data analytics plans

#### Scoring Process

We will develop an automated scoring and risk assessment system for trainees based on their current risk of mental health problems. The questionnaires completed by participants will be automatically scored immediately.

#### Databases

We will ensure that data collection and management are conducted rigorously and ethically, fully respecting the confidentiality and privacy of the information provided. Data will be stored in a secure, restricted-access database hosted on servers that implement appropriate security measures to protect sensitive participant information. Access to the database will be limited to authorized research personnel, all of whom will be bound by confidentiality and ethical regulations. The database will contain all information related to the scales used. Data will be stored and analyzed anonymously, ensuring that no individual participant can be identified from any published or disclosed results. Further sensitive information will be encrypted.

#### Statistical Analysis

We will estimate means, medians, interquartile ranges, and standard deviations for quantitative variables. We will first assess whether quantitative variables follow a normal distribution; if not, we will normalize them using z-scores for statistical contrasts. T-tests or ANOVAs will be used to compare quantitative variables between groups. For qualitative variables, we will estimate percentages and the prevalence of mental health problems. Differences in prevalence between groups will be compared using the chi-squared test. Given the longitudinal structure of MINDS, we will track changes over time in the presence of mental health problems using scales with temporal measurements, such as the PHQ-9, GAD-7, C-SSRS, and EDE-Q. We will model changes in symptoms to identify patterns and trajectories of onset over time using mixed linear models. All statistical analyses will be performed using open-source Python or R packages.

#### Power Analysis

##### Power for Differences in Prevalence

MINDS aims to enroll all undergraduates participating in medical internships and social service across all medical programs in Mexico, thereby creating a population-based study. We estimated the power to detect differences in the prevalence of mental health problems between sexes and between states.

A previous census of undergraduates participating in medical internships and social service estimated a total enrollment of 13,300 trainees. We conducted a power analysis to detect differences in prevalence ranging from 0% to 30%. Assuming a 1:1 male-to-female distribution, we found that we will be well powered to detect differences in prevalence exceeding 5.0% between the sexes. Assuming a 1:1 distribution across states, we also found that we will be well-powered to detect differences in prevalence greater than 10.0% between states (**Figure 3**).

**Figure 3.**
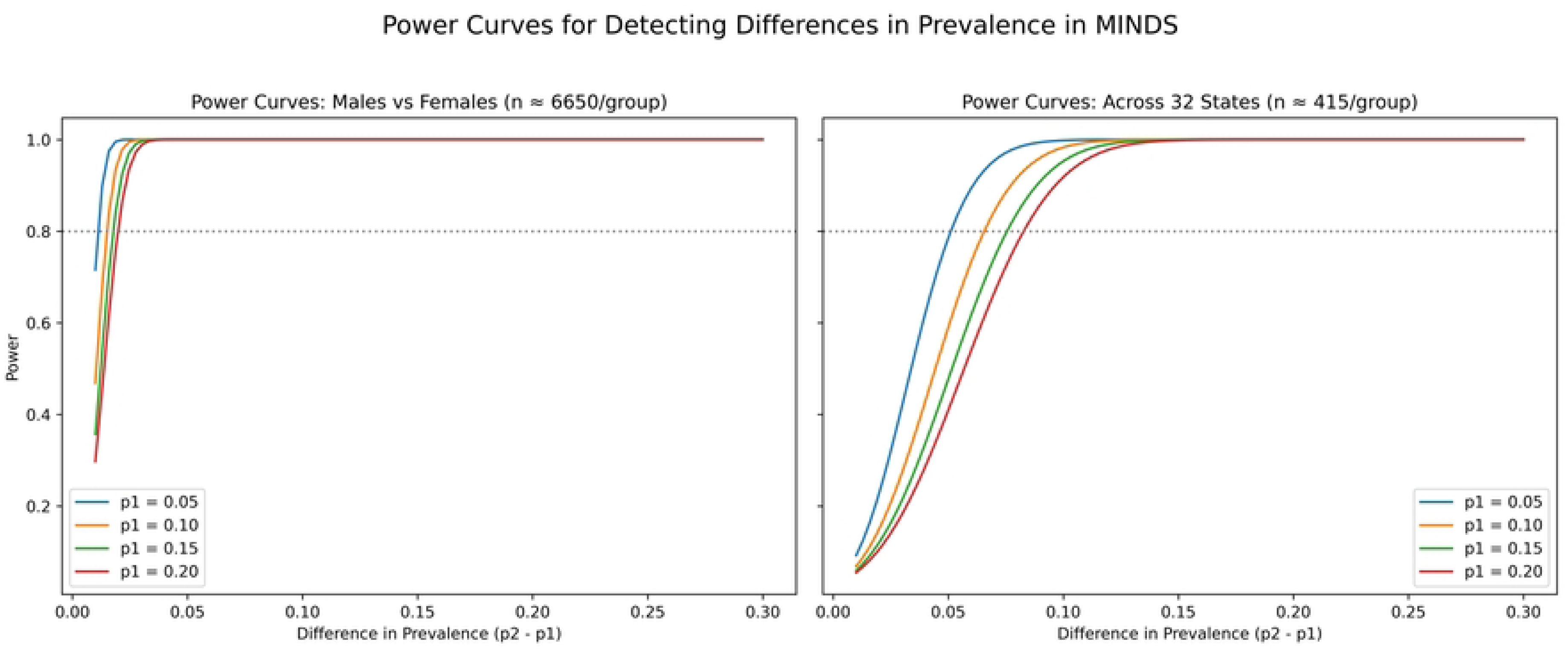
Power analysis for detecting differences in the prevalence of mental health problems between sexes and between states. The analysis was conducted assuming a total sample of 13,300 medical trainees, with an equal (1:1) distribution by sex and state. The figure illustrates the minimum detectable difference in prevalence at 80% power across varying baseline prevalence rates.

##### Power for Polygenic Risk Score (PRS) Analysis

We modeled our analysis using a well-powered genome-wide association study (GWAS) comprising 500,000 individuals as the training sample and up to 13,300 medical trainees as the target sample for PRS analysis. Our results indicate that genotyping 2,000 individuals would provide sufficient statistical power to detect a phenotypic variance explained by the PRS of 0.05% or greater (**Figure 4**).

**Figure 4.**
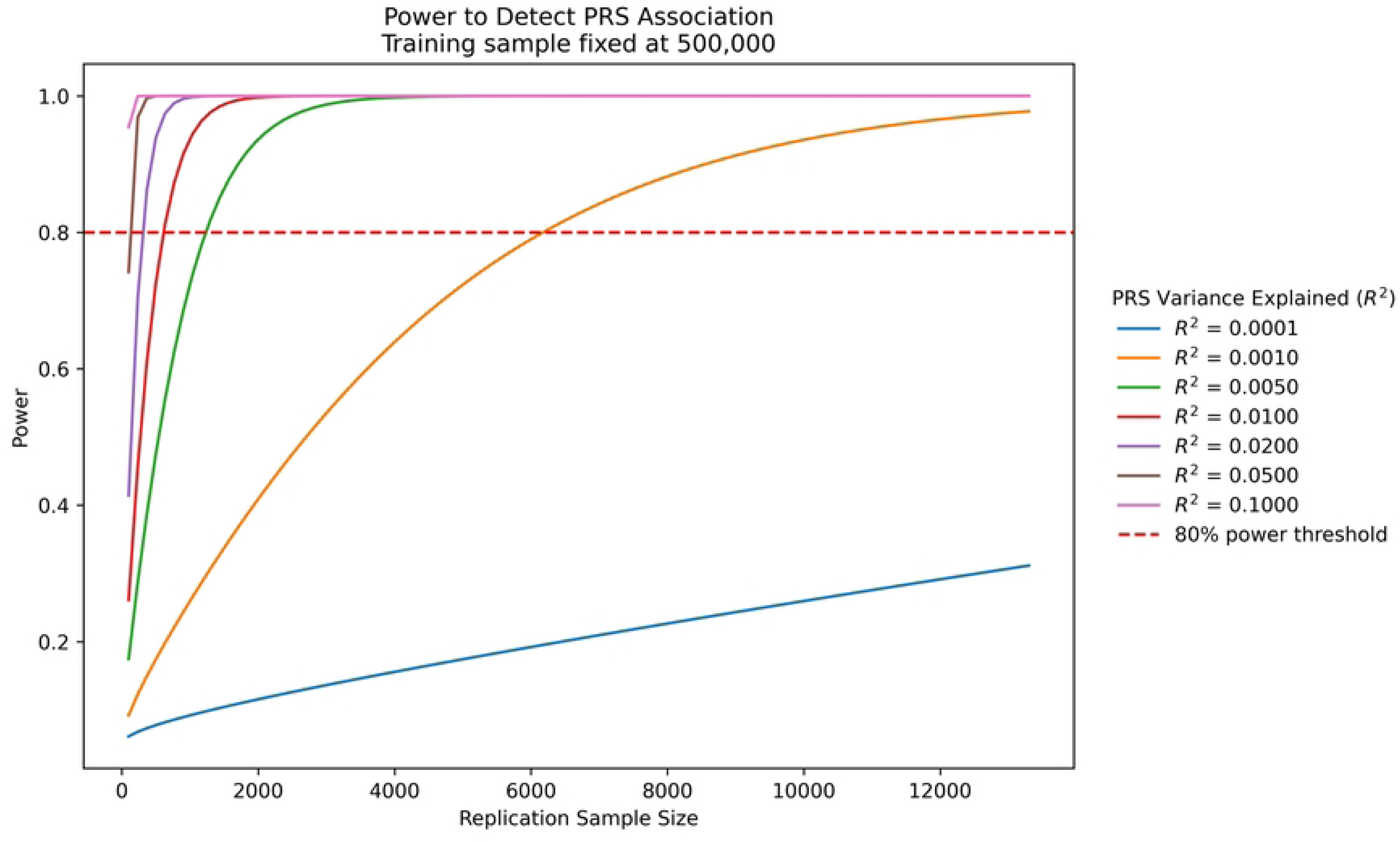
Sample size and power analysis for the polygenic risk score (PRS). We estimated the relationship between sample size and statistical power to detect phenotypic variance explained by the PRS, assuming a training GWAS of 500,000 individuals and up to 13,300 medical trainees in the target sample.

## Discussion

MINDS will address a critical and well-documented gap in trainee mental health care by combining systematic screening, automated risk stratification, and coordinated referrals across university and state health systems. MINDS, initiated by the Mexican Ministry of Health to enhance the well-being of medical trainees, recognizes the problem and raises awareness of the need to increase inter-institutional coordination to develop better strategies for medical trainees. Preliminary results of several studies on medical trainees show elevated rates of depression, anxiety, and suicidal ideation among Mexican medical students and trainees [27–29], pointing to the imperative need for developing programs such as MINDS.

The MINDS program builds upon a growing body of international evidence supporting scalable, preventive, and system-integrated approaches to mental health among medical trainees. For example, the Intern Health Study [30] in the United States (US) is a prospective annual cohort study that assesses the mental health of incoming US-based resident physicians in more than 150 programs. This program offers a valuable precedent for longitudinal monitoring of mood, stress, and biological data in medical trainees. MINDS mirrors other digital and telehealth initiatives that demonstrate the potential for confidential, scalable psychological support for medical students and trainees. For instance, the telemedicine service Apoyo Psicológico a Estudiantes de Medicina (SAPEM) in Spain offers a program to provide medical students with preventive, efficient mental health treatment. In Mexico, other programs with similar aims are being developed, such as the university psychological services, Espora Psicológica. This program provides evaluation and education of university students, targeted to generate training in mental health management. MINDS will not only advance mental health care for medical trainees but also foster a more open and supportive environment for discussing mental health within the physician community, addressing a well-known barrier that has historically limited physicians’ access to appropriate mental health care worldwide [31]. Together, MINDS is positioned as a timely, evidence-informed, and contextually grounded initiative with the potential to transform the mental health landscape for medical trainees in Mexico.

Additionally, MINDS will include a genomic sub-study, which will follow ethical governance and align with Mexican biorepository policies. The samples collected will be invaluable for future studies to understand better how genetic factors influence psychiatric and suicide risk, not only in medical trainees but in the whole Mexican society and ensure the high value of the collected specimens.

## Limitations

An explicit limitation of MINDS is the difficulty of maintaining follow-up once trainees complete their training period. Trainees frequently relocate, change contact information, and transition into heterogeneous residency programs or other employment, increasing the risk of follow-up and complicating longitudinal tracking. Besides, follow-up across residency programs introduces additional challenges. Residency programs are administratively and clinically distinct from undergraduate internship systems and are governed by different institutions and state-level arrangements. Establishing data-sharing agreements, ensuring consent for continued contact, and obtaining approval are necessary, but each requires a different approach. To advance the mental health of healthcare providers in Mexico, we will explore potential collaborations with major residency programs and national training bodies to facilitate follow-up care.

## Data Availability

No datasets were generated or analysed during the current study. All relevant data from this study will be made available upon study completion.

## Author contributions (CRediT taxonomy)

**Conceptualization:** Alma Delia Genis-Mendoza, José Jaime Martínez-Magaña

**Methodology:** Pamela Espinosa Méndez, Abril Téllez Buendía, Rocío Jurado Galicia, Carlos Alfonso Tovilla-Zárate, Thelma Beatriz Gonzales-Castro, José Jaime Martínez-Magaña

**Formal Analysis:** José Jaime Martínez-Magaña

**Investigation:** Pamela Espinosa Méndez, Abril Téllez Buendía, Rocío Jurado Galicia, Francisco Gutiérrez Rodríguez, Marta Catalina Pérez Gonzales

**Resources:** Humberto Nicolini, Carlos Alfonso Tovilla-Zárate, Thelma Beatriz Gonzales-Castro, Alma Delia Genis-Mendoza

**Writing – Original Draft:** Pamela Espinosa Méndez, José Jaime Martínez-Magaña

**Writing – Review & Editing:** All authors

**Visualization:** José Jaime Martínez-Magaña

## Acknowledgment

We extend our sincere gratitude to the educational and medical authorities whose support and collaboration were instrumental in advancing this project. Their willingness to contribute expertise, facilitate access to critical resources, and foster an environment conducive to research excellence greatly strengthened the development and implementation of our work. We are deeply appreciative of their commitment to improving scientific understanding and promoting the well-being of the communities we serve.

